# Wearable-Echo-FM: An ECG-echo foundation model for single lead electrocardiography

**DOI:** 10.1101/2025.06.10.25329163

**Authors:** Elizabeth Knight, Evangelos K Oikonomou, Arya Aminorroaya, Aline F Pedroso, Rohan Khera

## Abstract

Artificial intelligence (AI) models can now detect patterns of structural heart diseases (SHDs) from electrocardiograms (ECGs), though scaling them requires the broader use of single–lead ECGs that are now ubiquitous in wearable and portable devices. However, model development for these devices is limited by a lack of diagnostic labels for SHDs for wearable ECGs. Here, we present Wearable–Echo–FM, a foundation model that encodes single-lead ECGs with information from echocardiographic text reports. Using 274,057 single-lead ECG-echo pairs from 77,378 adults (2015–2019), we contrastively pre–trained convolutional neural network (CNN) and RoBERTa encoders. The ECG encoder was fine–tuned on a distinct progressively larger ECG set (250 to 250,260 ECGs) to detect different cardiac disorders (i) left–ventricular systolic dysfunction (LVSD), (ii) diastolic dysfunction, and (iii) a composite SHD. This was compared with a randomly initialized CNN, with both approaches evaluated in an independent held-out test set. With the full training set, Wearable–Echo–FM matched the baseline CNN (AUROC 0.894 vs 0.884 for LVSD; 0.849 vs 0.843 diastolic dysfunction; 0.887 vs 0.869 composite). With only 0.5% (~1000 ECGs) of data, it markedly outperformed baseline (0.855 vs 0.548; 0.819 vs 0.582; 0.863 vs 0.496, respectively). Contrastive pre–training of single-lead ECGs on echocardiographic text reduces label requirements for SHD screening on wearable and portable devices.

---

Structural heart diseases (SHD), including a range of myocardial and valvular abnormalities, represent modifiable conditions that pose a risk of heart failure (HF) and premature mortality.^1,2^ Early diagnosis is essential for the timely initiation of evidence-based therapies that can slow disease progression and reduce morbidity and mortality.^3,4,5,6,7^ Despite the availability of effective treatments, SHD remains frequently underdiagnosed, often only identified when patients present with overt symptoms.^5,4,8,9^ The reliance on resource-intensive cardiac imaging, specifically echocardiography, for diagnosing these conditions, limits early detection, particularly in low-resource settings.^10^

Recent advances in artificial intelligence (AI) have demonstrated the potential of electrocardiograms (ECGs) as screening tools for various cardiac conditions.^1,11,12,13,14^ Traditional 12-lead ECGs have been successfully leveraged for AI-based disease prediction; ^15,12,11^ however, single-lead ECGs, now integrated into widely available wearable and handheld devices, offer a more accessible alternative for large-scale screening.^15,16,14,17^ Early applications show the role of single-lead ECGs obtained from wearable and portable devices in identifying left ventricular systolic dysfunction (LVSD), suggesting that the 1-lead data might have sufficient information to capture SHDs.^14,1^ However, the development of AI models for SHD screening from single-lead ECGs remains constrained by the limited availability of labeled datasets, particularly for rare cardiac conditions.^18,19^ Moreover, development of models for individual diseases is resource-intensive, especially when limited labeled data are likely to be available for each condition.

To address these challenges, we propose an ECG-Echo foundation model (Wearable-Echo-FM) that learns shared representations between single-lead ECGs and structural cardiac phenotypes from unstructured echocardiographic reports. By leveraging contrastive pre-training and transfer learning, we sought to enhance the efficiency of AI models for SHD screening from 1-lead ECGs obtainable on wearable and portable devices. We hypothesize that our approach will enable more accurate detection of SHDs, even in settings with limited labeled data, thereby improving the accessibility and scalability of AI-based cardiovascular screening.

## Methods

### Study Overview

Based on data from the electronic health records (EHR) of the Yale New Haven Health System (YNHHS), which encompasses five hospitals and affiliated sites across Connecticut and Rhode Island, we developed a foundation model using transthoracic echocardiography (TTE) reports and concurrent single-lead ECGs. The study was approved by the Yale Institutional Review Board, which waived the need for informed consent as the research involved secondary analysis of existing data. Initially, we identified a subset of patients who had undergone both TTE and ECG within 30 days of each other between 2015 and 2018 for pretraining and contrastive learning, which created a foundation model we named Wearable-Echo-FM. We then set aside a temporally distinct cohort of patient data recorded between 2019 and 2023 for model fine-tuning and evaluation. We divided these 2019–2023 ECG-TTE pairs into three task-specific cohorts according to each structural heart disease (SHD) label described below. Each task’s cohort was used independently for training, validation, and testing to evaluate performance on that specific label. Subsequently, a temporally distinct and age-and sex-matched dataset from 2019-2023 was used to identify SHD conditions. We fine-tuned each model with varying amounts of data to compare the effectiveness of the wearable-echo-FM initialization (weights trained from the contrastive pretraining step) versus random initialization.

### Data Source and Study Population

Patient-level EHR data were collected from the YNHHS, encompassing a diverse patient population where patients were either seeking inpatient or outpatient care. Patients aged 18 or older with both a TTE and an ECG within 30 days of each other between 2015 and 2023 were included in the study. If multiple ECGs were recorded within 30 days of the TTE, a maximum of five ECGs were included per patient in the training set. We excluded patients who, at the time of ECG recording, had a history of cardiac procedures such as coronary artery bypass grafting, aortic or mitral valve procedure, left ventricular assist device implantation, heart transplant, alcohol septal ablation, or ventricular myectomy. Exclusion of these patients aimed to prevent an overrepresentation of individuals with frequent ECG recordings, who are also those who were unlikely to be candidates for screening using wearable and portable devices. The dataset included demographics (age, gender, ethnic group), ECG data such as the date of the ECG recording, and corresponding cardiac diagnoses on the TTE, as noted below.

### Study Outcome/Outcome Labels

Our study focused on 3 classification tasks using single-lead ECGs, each addressing critical markers of cardiac health. First, we predicted left ventricular ejection fraction (LVEF) ≤40%, a well-established threshold for defining LVSD. Second, we classified patients with left ventricular diastolic dysfunction, as determined by echocardiographic imaging. Lastly, we predicted a composite measure of SHDs. The composite SHD label was defined as the TTE-confirmed presence of LVSD, moderate or severe left-sided valvular diseases, and/or standardized left ventricular hypertrophy (sLVH). Moderate or severe left-sided valvular diseases were identified by the presence of any moderate or severe aortic regurgitation (AR), aortic stenosis (AS), mitral regurgitation (MR), or mitral stenosis (MS). We characterized sLVH by an interventricular septal diameter at end-diastole (IVSd) greater than 15mm with concomitant moderate or severe left ventricular diastolic dysfunction. By predicting myocardial and valvular abnormalities, systolic and diastolic dysfunction, as well as functional and structural cardiac parameters, we sought to explore the breadth of phenotypic information encoded within single-lead ECGs using the Wearable-Echo-FM model. With the model finetuning data (2019-2023), we identified separate cohorts of ECG-TTE pairs for each classification task: LVEF ≤40%, left ventricular diastolic dysfunction, and a composite of SHD. We used these task-specific datasets to fine-tune and evaluate the Wearable-Echo-FM model’s performance in predicting each clinical outcome through validation and testing. Details of the validation and test datasets are included in Supplementary Methods.

### Model Training

The study data were divided into two distinct subsets, which we split as pre-2019 data (2015-2018) for pretraining and contrastive learning purposes through a contrastive language-image pre-training (CLIP) framework,^20^ and used the temporally distinct subset of ECGs done during 2019-2023 for finetuning (figure 1).

**Figure 1:**
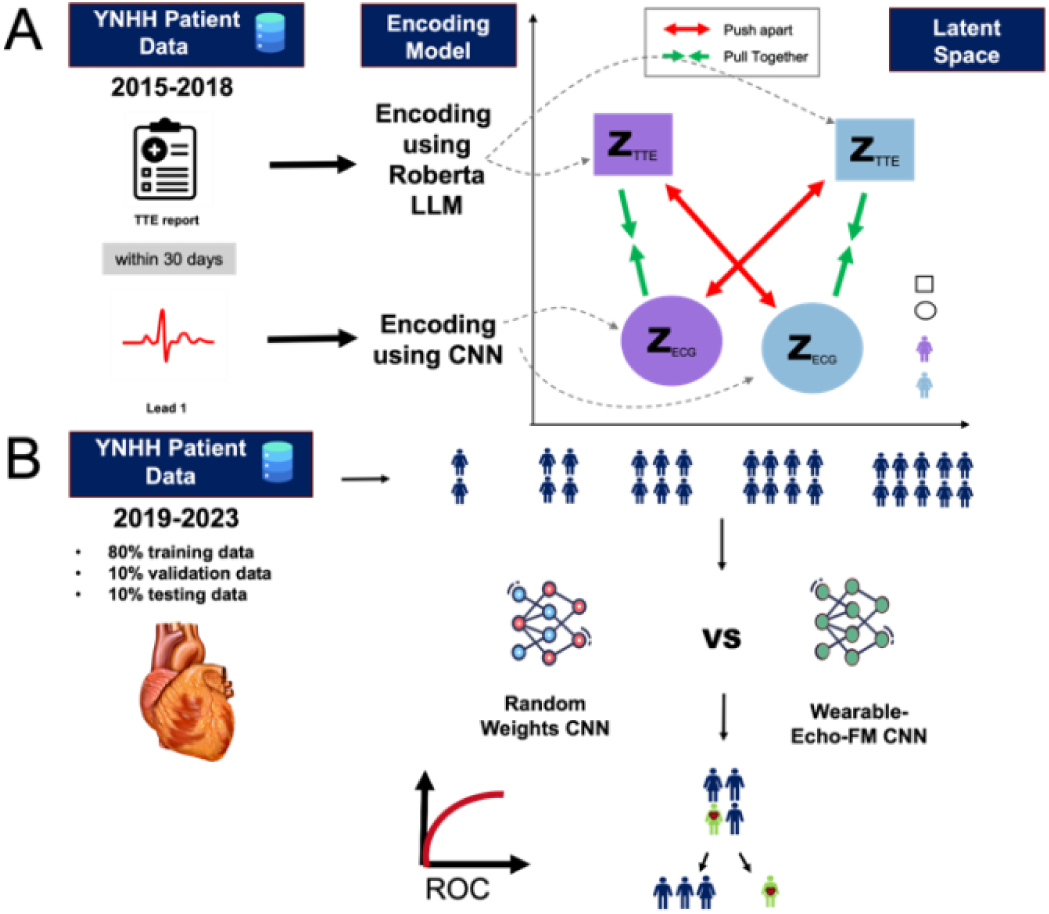
Overview of data flow and CLIP–style pre–training. Patient records from the YNNHS that contained paired TTE reports and ECGs acquired ≤ 30 days apart were split into 2015-2018 data (A) for contrastive pre–training and 2019–2023 data (B) for downstream fine–tuning and evaluation. Pannel A shows pre–training; a CNN encodes single–lead ECG signals while a RoBERTa–based encoder processes the corresponding TTE text. The two embeddings (Z_ECG_, Z_TTE_) are pulled together for matching pairs (green arrows) and pushed apart for non–matching pairs (red arrows), aligning both modalities in a shared latent space. The weights learned in this stage serve as initialization for subsequent fine–tuning on the temporally distinct cohort. Pannel B shows fine-tuning and testing; pre–trained weights are transferred to a distinct 2019–2023 cohort, split 80 / 10 / 10 % (train/validation/test) at the patient level (≤ 5 ECGs per subject in train; 1 in validation/test). Separate classifier heads are trained for three echo–derived labels (LVEF ≤ 40 %, LV diastolic dysfunction, and composite SHD) using cross–entropy with class weighting and early stopping (patience = 6). To test label–efficiency, the training set is randomly down–sampled to 0.1 %, 0.5 %, 1 %, 5 %, 20 %, 50 %, and 100 %. AUROC is used to measure classification ability. Abbreviations: YNNH, Yale New Haven Hospital. CNN, convolutional neural network. LLM, large language model. FM, Foundation Model. TTE, Transthoracic Echocardiogram. ECG, electrocardiogram.

### Model Architecture

We employed a seven–layer 1–D convolutional neural network (CNN) to encode the single–lead ECG waveform. This CNN is based on a previously described architecture by our group and with a base architecture with seven convolutional layers where each convolutional layer is followed by batch normalization, rectified linear unit (ReLU) activation, and max pooling, with the number of filters progressively increasing from 16 to 64 as the network deepens. The CNN incorporates dropout for regularization and terminates with a final embedding size of 512 (**Figure 3**)^21^. For processing textual data, we implement a transformer-based model inspired by Robustly Optimized BERT Pre-training Approach (RoBERTa), with 12 attention heads, 6 hidden layers, a maximum position embedding of 514 and vocabulary size of 1. We initially trained a ByteLevelBPETokenizer for this encoder and implemented the model with well-established hyperparameter values and architecture settings. Like the ECG encoder, the text encoder’s final embedding size is also 512. Both the ECG and text encoders project their outputs into a shared 512-dimensional embedding space, to enable contrastive learning.

The classification model was built upon the single lead encoder by including additional fully connected layers followed by a sigmoid activation with a dropout rate of 0.5 to prevent overfitting. Specifically, these fully connected layers consisted of two custom fully connected layer units, the first with 64 neurons and the second with 32 neurons. These were followed by a flattening operation and a final linear layer that mapped the 320 flattened features to 2 output neurons. A sigmoid activation was then applied to these two outputs, producing probability-like scores for binary classification.

### Pretraining

We initialized the weights of our single-lead ECG CNN using a CLIP-inspired approach that leveraged paired echocardiogram text data. We used masked language modeling to first pretrain the text encoder, a RoBERTa-based model, on TTE reports from the pre-2019 dataset to acclimate the model to better interpret Yale New Haven Hospital echocardiogram reports. To ensure sufficient textual content and quality, we filtered the data to include only reports with a string length of at least 100 characters, resulting in 87,342 reports used for pretraining. The median number of words per report was 256 (IQR, 198–310), indicating moderate variability in report lengths. We trained a Byte-Pair Encoding (BPE) tokenizer using the ByteLevelBPETokenizer, with a vocabulary size of 8,192 tokens and a minimum frequency threshold of 3 for token inclusion. The tokenizer was trained on the preprocessed reports, which were converted to lowercase and had dates and other identifying information removed to ensure anonymization.

The RoBERTa model was pretrained on TTE reports alone for 150 epochs with a learning rate of 10^−4^ and a weight decay of 0.01, using a masked language modeling probability of 0.15 to predict masked tokens in the input text. We used a batch size of 164 and employed the Adam optimizer for training. On a patient level, the data was randomly split into training and evaluation sets with a 90:10 ratio. In the training dataset, the median number of tokens per report was 260 with an IQR of [202–314], while the evaluation dataset had a median of 258 tokens (IQR [200–312]). This pretrained language model served as the textual encoder in our CLIP-inspired framework, facilitating the alignment of ECG signals with echocardiogram-derived textual features.

Next, a CNN was used in conjunction with the pre-trained RoBERTa language model to perform CLIP pretraining, where contrastive learning was employed to align the features extracted from the ECG data with the semantic information from the TTE reports. By training these components together, the goal was to develop pre-trained weights for the CNN that encapsulate rich, multimodal features. The model was trained with a batch size of 80, a cosine learning rate of 10^−4^, a weight decay of 10^−8^, for 200 epochs with 100 warm-up steps, and a maximum target length of 400. Early stopping was implemented with a patience of five epochs. As previously mentioned, the CLIP model used a projection dimension of 512 and a temperature of 1 and minimizes the contrastive loss (**Figure 1A**).

### Classification

Following pretraining, we transferred the model weights to ECG-report pairs acquired between 2019 and 2023 in a distinct set of patients from those included in the contrastive pretraining. We split the dataset of ECG-TTE pairs randomly by patient ID into an 80-10-10 split where 80% of the data was allocated for training, 10% was allocated for evaluation to enable early stopping, and 10% was used for the final test set. The training dataset was further filtered to include a maximum of five ECGs per individual, while the validation and testing datasets were filtered to include only one ECG per person. Echocardiography labels (LVEF ≤ 40%, left ventricular diastolic dysfunction, or composite SHD) were derived from paired TTEs (**figure 1B**).

To assess performance with varying amounts of labeled data, we fine-tuned separate models for each clinical label using progressively smaller random fractions from the model fine-tuning cohort. We sampled 0.1%, 0.5%, 1%, 5%, 20%, 50%, and 100% of the available training data and trained the model from the same initialized weights under each condition. For each fraction, we randomly selected the corresponding percentage of ECG-report pairs while maintaining the same validation and test splits for consistency. Since some labels had fewer available examples, the exact sample size for each fraction varied across tasks. We trained each model with early stopping and patience of 6 epochs to prevent overfitting, using cross-entropy loss with class weights to account for label imbalance. All other hyperparameters (learning rate, batch size, optimizer) were the same as in the base fine-tuning setup. The final model performance was reported on the test set (**Figure 1B**).

### Statistical Analysis

We summarized demographic characteristics using medians with interquartile ranges (IQR) for continuous variables and counts with percentages for categorical variables. The model’s testing performance for each training data subset was evaluated by calculating the area under the receiver operating characteristic curve (AUROC). To obtain the 95% confidence interval for the AUROC, we employ bootstrapping. All tests were two-sided, and statistical significance was set at an alpha level of 0.05. The analyses were performed using Python 3.9 with NumPy, Pandas and Scikit-learn libraries.

## RESULTS

### Study Population

We included 524,317 ECG-TTE pairs from patients who underwent both procedures within 30 days of each other between 2015 and 2023. These pairs were divided into distinct cohorts for model development and finetuning. The pretraining cohort, used for the development of Wearable Echo-FM, included 274,057 ECG-TTE pairs from 77,378 unique individuals. For task specific model finetuning and testing, we included three post-2019 cohorts: LVEF ≤ 40% (250,260 ECGs from 95,388 individuals), left ventricular diastolic dysfunction (123,306 ECGs from 59,831 individuals), and a SHD (132,310 ECGs from 58,815 individuals). Additionally, we had separate validation and test sets for each condition.

In the pretraining cohort, patients had a median age of 68.2 years (IQR 57.0, 78.5) at the time of the ECG and 117,966 (43.0%) women. The cohort included 196,524 (71.7%) White, 41,506 (15.1%) Black, 3,548 (1.3%) Asian, 23,549 (8.6%) Hispanic and 2,440 (.9%) unknown individuals. The model finetuning cohorts had a median age of 69.6 years (IQR 58.4, 79.5), with 116014 (46.4%) women. The population included 172,190 (68.8%) White, 32,921 (13.2%) Black, 3,587 (1.4%) Asian, 21,423 (8.6%) Hispanic and 14,048 (5.6%) Unknown. The proportions of individual labels remained relatively consistent across the validation and test sets.

### Pretraining results

During contrastive pre–training, the ECG and text encoders were optimized jointly for 200 epochs with a batch size of 80, Adam (β_1_ = 0.9, β_2_ = 0.999), and a cosine learning–rate schedule. The cross–modal contrastive loss fell from 4.19 after the first epoch to 0.012 by epoch 200 when pretraining was set to stop.

### Performance in held-out test set

When trained on the full model finetuning cohort, our Wearable-Echo-FM model achieved comparable performance to the standard CNN with randomly initialized weights. For detecting LVEF ≤ 40%, the Wearable-Echo-FM model achieved an AUROC of 0.894 (95% CI: 0.888-0.900) compared with 0.884 (95% CI: 0.878-0.890) for the standard CNN. For LV diastolic dysfunction, the AUROCs were 0.849 (95% CI: 0.842-0.856) and 0.843 (95% CI: 0.836-0.850) respectively. For the composite of SHD outcome, the Wearable-Echo-FM model achieved an AUROC of 0.887 (95% CI: 0.881-0.893) versus 0.869 (95% CI: 0.863-0.875) for the standard CNN (**Table 2**).

**Table 1:**
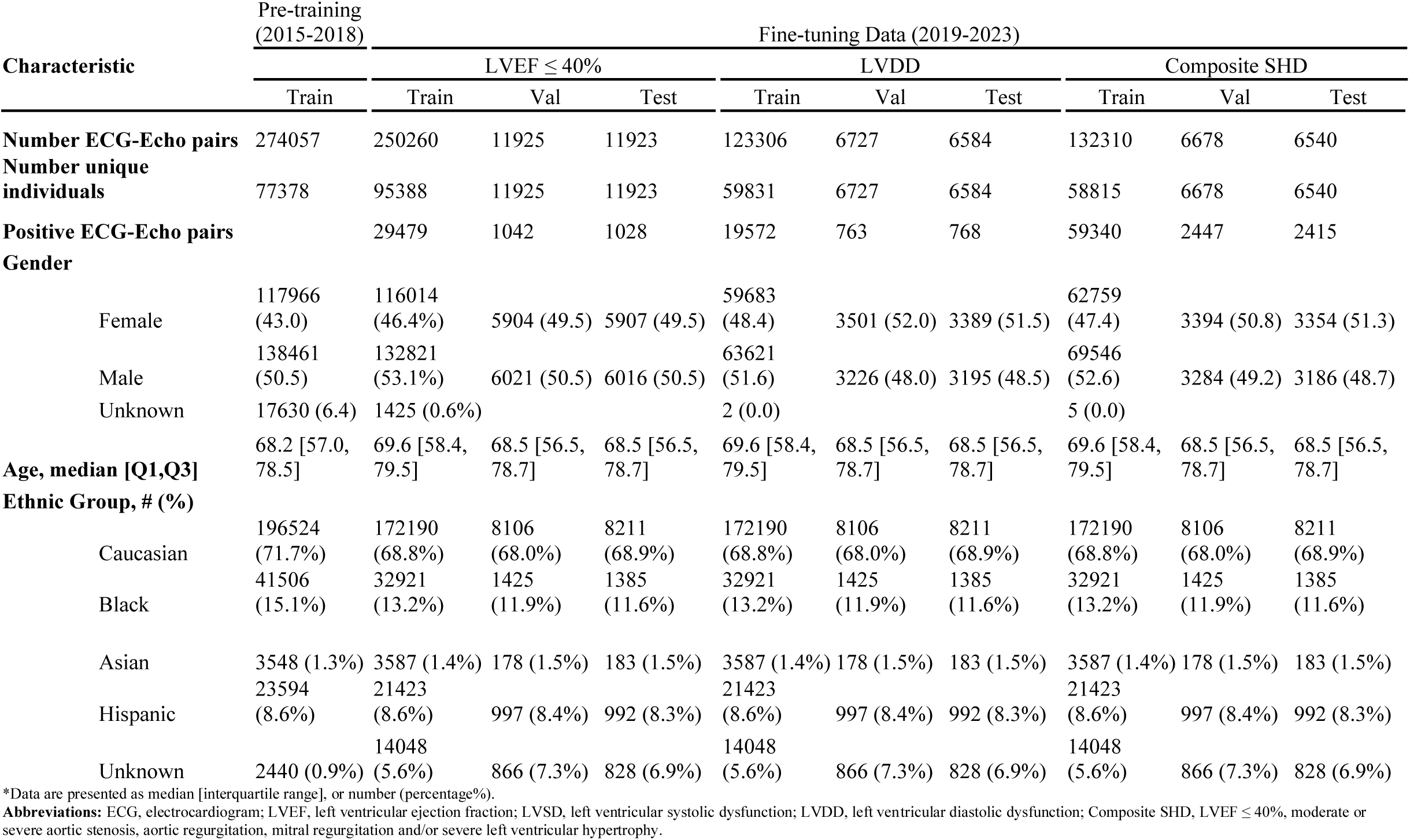
Demographic and clinical characteristics of the model development population.

**Table 2:** Performance metrics for detecting LVEF, LVDD and SHDs in the held-out test set.

| Label | Percentage of Training Data | Test AUROC |  |
| --- | --- | --- | --- |
|  |  | Wearable-Echo-FM | Standard Model |
| LVEF $\leq 40$ | 0.001 | 0.841 [0.828, 0.853] | 0.491 [0.473, 0.51] |
|  | 0.005 | 0.854 [0.842, 0.865] | 0.548 [0.529, 0.567] |
|  | 0.01 | 0.863 [0.851, 0.874] | 0.841 [0.828, 0.853] |
|  | 0.05 | 0.876 [0.865, 0.886] | 0.857 [0.845, 0.868] |
|  | 0.2 | 0.883 [0.872, 0.893] | 0.874 [0.862, 0.884] |
|  | 0.5 | 0.887 [0.876, 0.896] | 0.88 [0.869, 0.89] |
|  | 1 | 0.894 [0.884, 0.903] | 0.884 [0.874, 0.894] |
| LVDD | 0.001 | 0.801 [0.775, 0.823] | 0.51 [0.477, 0.542] |
|  | 0.005 | 0.819 [0.795, 0.839] | 0.582 [0.549, 0.615] |
|  | 0.01 | 0.806 [0.781, 0.827] | 0.782 [0.758, 0.805] |
|  | 0.05 | 0.843 [0.822, 0.861] | 0.804 [0.779, 0.827] |
|  | 0.2 | 0.843 [0.821, 0.861] | 0.838 [0.815, 0.856] |
|  | 0.5 | 0.851 [0.83, 0.869] | 0.851 [0.831, 0.869] |
|  | 1 | 0.849 [0.828, 0.866] | 0.843 [0.822, 0.862] |
| Composite SHD | 0.001 | 0.486 [0.466, 0.505] | 0.503 [0.482, 0.524] |
|  | 0.005 | 0.864 [0.851, 0.875] | 0.496 [0.477, 0.516] |
|  | 0.01 | 0.865 [0.853, 0.876] | 0.831 [0.817, 0.843] |
|  | 0.05 | 0.862 [0.85, 0.873] | 0.853 [0.84, 0.864] |
|  | 0.2 | 0.878 [0.867, 0.889] | 0.859 [0.846, 0.87] |
|  | 0.5 | 0.873 [0.862, 0.883] | 0.868 [0.856, 0.879] |
|  | 1 | 0.887 [0.876, 0.896] | 0.869 [0.858, 0.88] |

\*Data are presented as AUROC, 95% confidence interval
**Abbreviations:** LVEF, left ventricular ejection fraction, LVDD, left ventricular diastolic dysfunction; Composite SHD, LVEF $\leq 40\%$ , moderate or severe aortic stenosis, aortic regurgitation, mitral regurgitation and/or severe left ventricular hypertrophy. AUROC, area under the receiver operating characteristic curve. Wearable-Echo-FM, pretrained model with weights initialized from pretraining step. Standard Model, with random weights initialized.

The model performance was dependent on the volume of training data for both standard and Wearable-Echo-FM. However, the Wearable-Echo-FM model consistently outperformed the standard CNN, with the performance gap widening as data became scarcer (**Figure 2**). In experiments using only 0.5% of the training data (approximately 1000 ECGs), the pretrained Wearable-Echo-FM model significantly outperformed the standard model across all tasks. For detecting LVEF ≤ 40%, the Wearable-Echo-FM model achieved an AUROC of 0.855 (95% CI: 0.848-0.862) compared with 0.548 (95% CI: 0.539-0.557) for the standard model. For left ventricular diastolic dysfunction detection, the AUROCs were 0.819 (95% CI: 0.811-0.827) compared with 0.582 (95% CI: 0.573-0.591) for the standard model and for composite SHD, the Wearable-Echo-FM model achieved an AUROC of 0.864 (95% CI: 0.851-0.875) versus 0.496 (95% CI: 0.477-0.516) for the standard model (**Table 2**).

**Figure 2:**
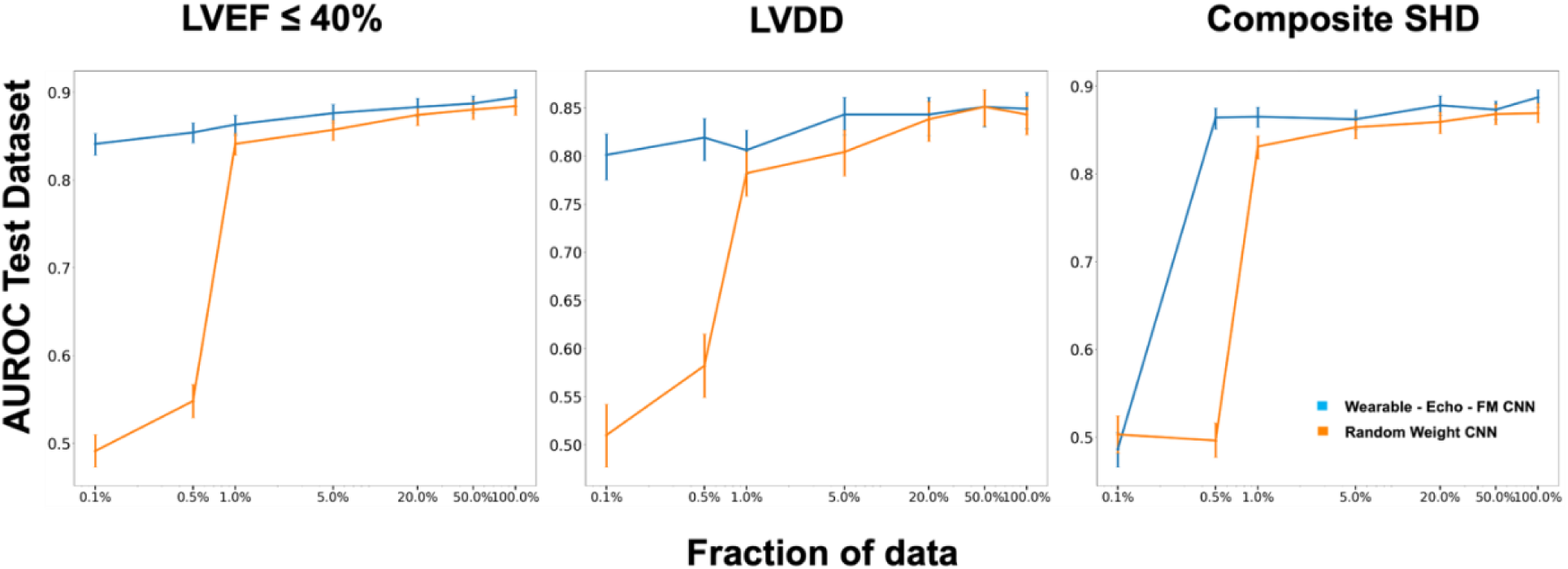
Model performance across different fractions of data for LVEF ≤40, LVDD, and the composite outcome. Test–set AUROC (mean ± 95 % CI) for detecting LVEF ≤ 40 %, LV diastolic dysfunction, and composite SHD as a function of the fraction of labeled training data used for fine–tuning (0.1 % to 100 %). The pre–trained Wearable–Echo–FM shown in blue and the randomly initialized CNN is shown in orange. Abbreviations: LVEF, left ventricular ejection fraction, LVDD, left ventricular diastolic dysfunction; Composite SHD, LVEF ≤ 40%, moderate or severe aortic stenosis, aortic regurgitation, mitral regurgitation and/or severe left ventricular hypertrophy. AUROC, area under the receiver operating characteristic curve. Wearable-Echo-FM, pretrained model with weights initialized from pretraining step. Standard Model, with random weights initialized.

**Figure 3:**
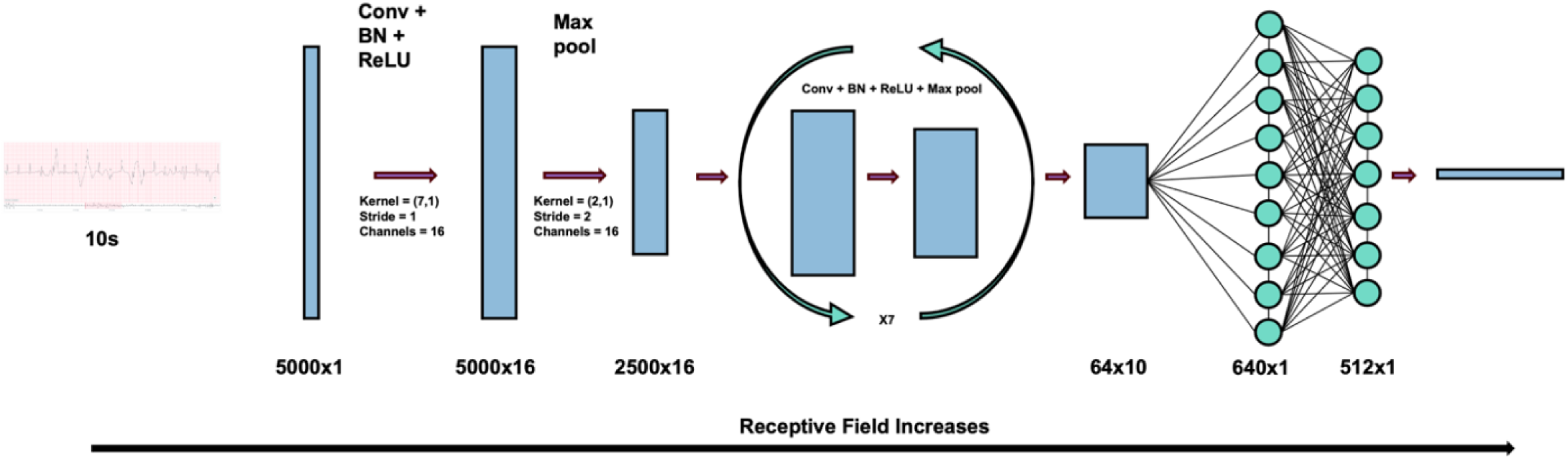
Signal encoder architecture optimized for embedding used in contrastive framework. The seven layer 1D CNN model encoded the 10s single lead ECGs. Each convolutional block consists of (Conv, BatchNorm, ReLU, MaxPool), with kernel sizes tapering from 7 to 3 and filters doubling every other layer (16, 32, 64) to capture both broad and fine–grained temporal features. Max–pooling strides of 2 and 4 progressively down–sample the 10s input while expanding the receptive field. The final convolutional output is flattened and passed through two FC layers (64 and 32 units) with 0.5 dropout, producing a 512–dimensional ECG embedding that feeds the contrastive loss. Abbreviations: CNN, convolutional neural network; ECG, electrocardiogram; Conv, convolution; BatchNorm, Batch normalization; ReLU, Rectified linear unit activation function; Max-pool, maximum pooling layer. FC, fully connected (dense) layer; L2-normalization, Euclidean (L2) vector norm;

## DISCUSSION

In this study, we introduce Wearable-Echo-FM, a foundation model optimized for screening SHDs from single-lead ECGs that leverages joint embeddings of ECG signals and paired echocardiographic reports. We demonstrate that the model enables efficient development of downstream applications, achieving superior performance relative to standard training techniques, especially when there are a limited number of cases. Notably, these findings were robust across analyses of various individual SHDs and a composite SHD measure, spanning features of systolic function, diastolic function, valvular abnormalities, and abnormal LV hypertrophy. These findings highlight Wearable-Echo-FM as a powerful model that can boost our ability to screen for SHD from single-lead ECGs increasingly available from wearable and portable devices in the community.

The study builds upon existing literature in developing deep learning models to enhance the diagnostic capabilities of ubiquitous wearable devices.^1,21^ Prior work has focused on two key domains. The first of which represents a series of tools built to detect observable rhythm and conduction disorders.^17^ Single-lead ECG sensors, integrated into consumer devices (such as watches or wristbands), can enable large-scale community-based screening and monitoring for conditions such as atrial fibrillation and bundle branch blocks.^5^ Advances in electrode placement and signal quality have allowed this technology to produce reliable single-lead data and detect straightforward, clinically observable patterns that have been validated.^22^ The second represents tools that identify signatures of a common structural heart disorder, LVSD, a “hidden” feature not identifiable by experts.^1,21^ In a prospective study in 2022 Attia et. al. at the Mayo Clinic demonstrated that an AI-ECG model using single-lead ECGs from smartwatches could detect LVSD with an area under the receiving operating characteristic (ROC) curve of approximately 0.88 in real-world settings.^23^ Likewise, our group showed that deep-learning models can robustly identify LVSD from single-lead ECG signals adapted for portable and wearable devices, even under highly noisy conditions.^21,1^ Building on these advances, our foundation model approach further broadens single-lead ECG–based screening to a broader range of SHDs without requiring extensive labeled datasets.

By leveraging single-lead ECGs readily obtainable from wearable and handheld devices, our framework enables broader community-level screening for SHDs.^24^ Early detection of these structural disorders, which often remain underdiagnosed until clinical symptoms arise, enables timely interventions that can substantially reduce morbidity, mortality, and healthcare costs.^10,7^ Through harnessing the deeper structural insights of echocardiographic findings, our approach provides an adaptable model that can be applied to underrepresented conditions where large, fully labeled datasets are scarce. As wearable ECG technology continues to be increasingly adopted, this foundation model can be fine-tuned for additional structural and functional cardiac abnormalities This adaptability broadens its clinical utility and offers a scalable, cost-effective solution for cardiac screening, particularly in resource-limited settings.

An important observation from our experiments is that the Wearable-Echo-FM model consistently outperformed the standard, randomly initialized CNN across a broad spectrum of training set sizes. For instance, when the model was fine-tuned with only 0.5% of the available data (roughly 1000 ECGs), the Wearable-Echo-FM still demonstrated robust detection capabilities for LVEF ≤40%, left ventricular diastolic dysfunction, and composite SHD, while the standard model’s performance degraded substantially. Even at higher fractions of the training data, the pre-trained model provided incremental gains, which suggests that its prior exposure to echocardiographic text and ECG pairings conferred possible advantage. Because of its training on a broad array of echocardiographic findings, Wearable-Echo-FM captures SHD features even under conditions of extremely limited labeled data.

This study has certain limitations that merit consideration. First, while the model was trained on single-lead signals extracted from clinical 12-lead ECGs rather than directly from consumer-grade wearables, prior work, including the prospective smartwatch study by Attia et al., has shown that single-lead algorithms for detecting LVSD retain strong performance in real-world wearable ECGs. These findings suggest that our model may similarly generalize to real-world wearable ECG data, though direct validation is still needed. Second, the model was evaluated based on SHD phenotypes defined by TTE, and its applicability to other cardiac conditions remains unexplored. Lastly, the model was developed within a single health system. However, the development cohort from the YNHHS is one of the most racially and ethnically diverse in the US and closely mirrors the national demographic distribution, supporting the potential generalizability of our findings across diverse populations. The inherent variability of echocardiographic measurements in clinical reads could introduce label noise and may affect reported performance. Prospective studies in broader community settings, ideally with diverse device manufacturers, are essential to confirm the scalability and long-term utility of our approach for widespread SHD screening.

## Conclusion

An ECG-Echo foundation model that learns shared embeddings between 1-lead ECGs and complex structural cardiac phenotypes recorded on unstructured echocardiographic reports enhances the efficiency of developing AI algorithms for SHD screening on wearable and portable devices.

## Data availability

The dataset cannot be made publicly available because they are electronic health records, and sharing this data externally without proper consent could compromise patient privacy and would violate the Institutional Review Board approval for the study.

## Computer code

The code for the study is available from the authors upon request.

## Acknowledgments

Dr. Khera was supported by the National Institutes of Health (under awards R01AG089981, R01HL167858, and K23HL153775) and the Doris Duke Charitable Foundation (under award 2022060). The funders had no role in the design and conduct of the study.

## Author Contributions

R.K. conceived and designed the study and accessed the data. E.K., A.A., E.K.O. and R.K. developed the model. E.K., E.K.O, A.A. and R.K. performed the statistical analyses. E.K.O., A.F.P. and E.K. and R.K. drafted the manuscript. All authors reviewed the study design, provided critical feedback, and approved the final version of the manuscript. R.K. supervised the project, secured funding, and is the guarantor.

## Materials & Correspondence

Rohan Khera, MD, MS

195 Church Street, 6^th^ Floor, New Haven, CT 06510

